# A revised mathematical model of pre-diagnostic glioma growth incorporating vascularization and tumor mutational burden

**DOI:** 10.1101/2024.01.08.24300708

**Authors:** Rishab K. Jain

## Abstract

Gliomas, a type of brain tumor, have become increasingly important in oncology as they are difficult to treat due to their location deep in the brain. While some research has been done on spatiotemporal prediction of future glioma growth—something that can aid in surgical resections of gliomas once a patient has been diagnosed—modeling efforts for pre-diagnostic gliomas remain limited. The lack of retrospective, pre-diagnostic data makes this a challenging task; yet, pre-symptomatic serum glucose levels in patients have been shown to have a relationship with the emergence of gliomas, motivating this area of research. In 2015, Sturrock et al. presented an ordinary differential equation model of pre-diagnostic glioma growth that describes glioma-glucose-immune interactions. This report reproduces the major findings of Sturrock et al., revising their model to incorporate more biological phenomena—namely vascularization and mutational burden—testing additional medically relevant patient scenarios, and providing an extended discussion on the implications of the model. *In-silico* simulations performed in this report provide further insight into models describing glioma-glucose-immune interactions, and how they can be expanded to incorporate physiologically relevant features. Future work is necessary to refine model parameters and validate predictions with the limited, albeit steadily growing, amount of longitudinal patient data.

## 1. Background and Context

Gliomas are a type of brain tumor that are known to be particularly difficult to treat, due to their location deep in the brain and their anisotropic growth. As discussed by Sturrock et al. (2015) in their review, while significant efforts have been directed towards studying the spatiotemporal growth of gliomas towards personalized treatment, there has been less attention on pre-diagnostic gliomas. These tumors—especially the glioblastoma subtype—are deadly, with high fatality rates, motivating research in this area. Pre-diagnostic gliomas refer to tumors that had existed, undetected often due to asymptomaticity, before the diagnosis and treatment for them was initiated. There is a lack of detailed information about pre-diagnostic gliomas due to their undetected nature and the difficulty in retrospectively collecting clinical data for such tumors. Despite this lack of data, pre-diagnostic gliomas have been associated with elevated serum glucose levels in patients, indicating that the metabolic status of these patients may have an influence on the emergence of gliomas (Edlinger et al., 2012; Flavahan et al., 2013). This has motivated researchers, such as Sturrock et al., to develop mathematical models describing preceding diagnosis in order to gain a better understanding of glioma-glucose interactions and to potentially facilitate earlier diagnoses and screening efforts.

The immune system plays a large role in fending off tumor growth, potentially providing a link between glucose metabolism and glioma growth. As immune response requires energy, an increase of serum glucose is key for providing the energy needed for the immune system (Sturrock et al., 2015). In order for immune cells to jump from rest to an active state, they use the energy from glucose metabolism. Thus, glucose levels are expected to play a role in glioma growth, which is said to be inhibited by the immune system. However, this relationship is not well understood, and there is further complexity that needs to be taken into account such as the local and tumor-specific environment within gliomas and the influence of pre-existing mutations on tumor growth and immune-avoidance. Immune cells can recognize and kill tumor cells that express certain antigens or markers on their surface. However, some tumor cells can escape the immune recognition by losing or altering these antigens or markers, or by creating an immunosuppressive microenvironment. Hence, there is an interest in measuring tumor mutational burden—specifically, understanding how and when a tumor accumulates mutations that allow it to evade the immune system (Wilkie et al., 2016).

As Sturrock et al. (2015) acknowledge, when tumors are vascular, they become more resistant to immune response. Specifically, avascular tumors are significantly inhibited by tumor-infiltrating cytotoxic lymphocytes. This highlights the importance of considering vascularization when trying to understand the interactions between glioma growth and the immune response. If immune response is high, but the tumor has not established a vascular network, then the immune response is more likely to be effective since there is limited nutrient supply and the tumor will be inhibited and its growth delayed (Yuan et al., 1996). Yuan et al. specifically investigate vascular permeability and vascular growth factor. These parameters depend on both tumor area in contact with vascular endothelium and the number of functional vessels. Angiogenesis—this process of blood vessel formation—thereby influences tumor growth. The hypothesis is that if the tumor is vascularized, i.e. its growth is further supported by a continuous supply of nutrients, then it is more likely to survive and continue to grow despite the presence of an immune response. Additionally, the immune response to the glioma—which is typically anti-tumor—may be diminished due to the presence of vascularization.

Sturrock et al.’s model utilizes ordinary differential equations, which differs from recent computational and partial differential equation approaches for tumor growth predictions (Jain et al., 2022; Lipkova et al., 2019). This is relevant—the Sturrock et al. mathematical model, while simplifying to a raw biomass calculation of the tumor, allows one to understand the effects of glucose and immune activity, making it mathematically and computationally simpler to model. It is a more general approach which suggests that it could still capture other processes, such as angiogenesis and mutational burden, which are crucial components of gliomas. This report will first reproduce the Sturrock et al. model, detailing challenges and limitations encountered. Next, revisions to the model’s components and parameters will be made to incorporate more biological phenomena around gliomas. A new, medically relevant test case regarding variable glucose inputs is laid out, and its results are discussed in-depth. Finally, a discussion is provided, along with directions for future works.

## 2. The Mathematical Model

The Sturrock et al. system of differential equations, describing glioma-glucose-immune interactions, is used here as the basis for further developments and testing. This model was based on the assumption that the pre-diagnostic glioma is an initial, small mass of tissue, and glucose is the primary driver for growth that is required for gliomas to advance towards a clinically diagnosable state. The schema of the original system of equations, as presented in Sturrock et al., is provided in 2.1, with definitions of model parameters. This system is simulated in 2.2, with the three biologically relevant steady states. In 2.3, the effect of increasing glucose intake is reproduced, and the tunability of the glucose intake function is investigated. Finally, some additional quantitative verification is performed in 2.4 with respect to bifurcations caused by a sensitive parameter.

### 2.1 Model Explanation and Parameters

The original Sturrock et al. model of pre-diagnostic glioma growth is the following system of differential equations, following mass-action kinetics, describing glioma-glucose-immune interactions:

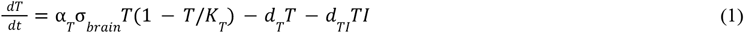

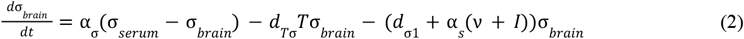

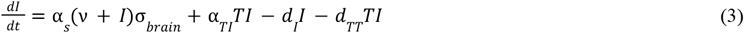

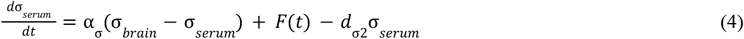

with initial conditions *T*(0) = *T*_0_ ; σ_*brain*_ (0) = σ_*brain*,0_, *I*(0) = *I*_0_, σ_*serum*_ (0) = σ_*serum*,0_ which must be four positive constants (negative values are not biologically relevant). *F(t)* is defined by:

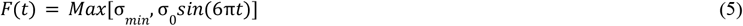

a variable function representing glucose intake.

Equation 1 describes the tumor’s growth, describing how tumor cells replicate and proliferate, while also being influenced by immune response. Equation 2 describes how glucose moves between serum and the brain, while also being influenced by, separately, the tumor and brain’s consumption. Equation 3 describes the interactions between tumor-tissue and immune cells, including the dynamics of their natural production. Equation 4 finally describes the interactions of serum glucose, influenced by glucose intake *F(t)*, as well as natural consumption. The parameters are given by Sturrock et al. in their original paper and specified in Table 1:

**Table 1.** Parameters for differential equation system, taken from Sturrock et al. (2015)

| Parameter | Description | Range (unit) |
| --- | --- | --- |
| $\alpha_T$ | Growth rate of glioma | $1.575 \text{ (ml}^2 \text{ g}^{-2} \text{ day}^{-1}\text{)}$ |
| $K_T$ | Carrying capacity of glioma | $2 \text{ (g/ml)}$ |
| $d_{TI}$ | Decay rate of glioma due to immune response | $0.072 \text{ (day}^{-1}\text{)}$ |
| $\alpha_{TI}$ | Recruitment rate of immune systems cells due to glioma | $3 \times 10^{-4} \text{ (day}^{-1}\text{)}$ |
| $d_T$ | Natural decay rate of glioma | $1 \times 10^{-4} \text{ (day}^{-1}\text{)}$ |
| $d_I$ | Natural decay rate of immune system cells | $0.01 \text{ (day}^{-1}\text{)}$ |
| $\alpha_s$ | Immune system cell recruitment rate | $0.7 \text{ (day}^{-1}\text{)}$ |
| $\nu$ | Baseline immune system cell production rate | $0.7 \text{ (day}^{-1}\text{)}$ |
| $d_{T\sigma}$ | Glucose consumption rate by glioma | $1 \text{ (day}^{-1}\text{)}$ |
| $\alpha_\sigma$ | Transfer rate of glucose from serum to brain | $20.0 \text{ (day}^{-1}\text{)}$ |
| $\sigma_{min}$ | Minimum glucose intake rate to serum | $8.00 \times 10^{-4} \text{ (g/ml)}$ |
| $\sigma_0$ | Maximum variation in glucose intake rate | $1.6 \times 10^{-3} \text{ (g/ml)}$ |
| $d_\sigma$ | Glucose consumption in brain by healthy cells | $0.01 \text{ (day}^{-1}\text{)}$ |
| $d_{TT}$ | Rate of glioma cells killing immune cells | $0.72 \text{ (day}^{-1}\text{)}$ |

Sturrock et al. assume that the glioma exhibits logistic growth and has a carrying capacity *K*_*T*_. Whilst being influenced by parameter α_*T*_, the glioma is also affected by glucose in the brain σ_*brain*_ . The tumor undergoes apoptosis and immune response which degrades its size. For both serum and brain glucose levels, they use an exchange term which describes transfer of glucose from the serum to the brain and vice versa, along with natural consumptions of glucose. The immune system activity is modeled based on cell production—which is influenced by glucose levels in the brain—and death caused by natural degradation and in interactions with the glioma.

### 2.2 Reproducing Model Steady-States

Based on their system, Sturrock et al. consider three equilibrium points or steady states^1^:

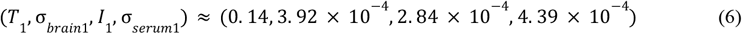

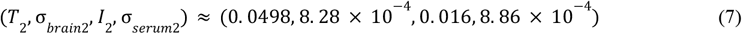

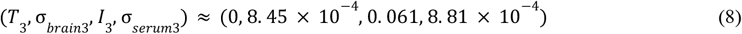

with conditions in units of g/ml where one gram contains approximately 10^9^ cells; glioma cells have a volume of approximately 10^−8^ ml. However, there appears to be several typos made in their establishment of these steady states, and they point to a later section which includes a discussion of all steady states. There, Equation 6 appears to be replaced with:

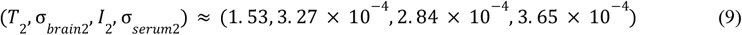

There are three steady states they describe: healthy patient/no tumor (Equation 8), small/dormant tumor (Equation 7), and large/aggressive tumor (Equation 6 and 9). Upon simulating steady states defined by Equation 6 and 9, there is a quite large difference between the two, with Equation 6 demonstrating sufficient tumor growth as shown in Figure 1, and Equation 9 showing a relatively stable (but large) tumor as shown in Figure 2 that is closer to the carrying capacity of 2 g/ml. Thus, it is clear that Equation 9 is representative of the large/aggressive tumor, and this is an error in their manuscript.

**Figure 1.**
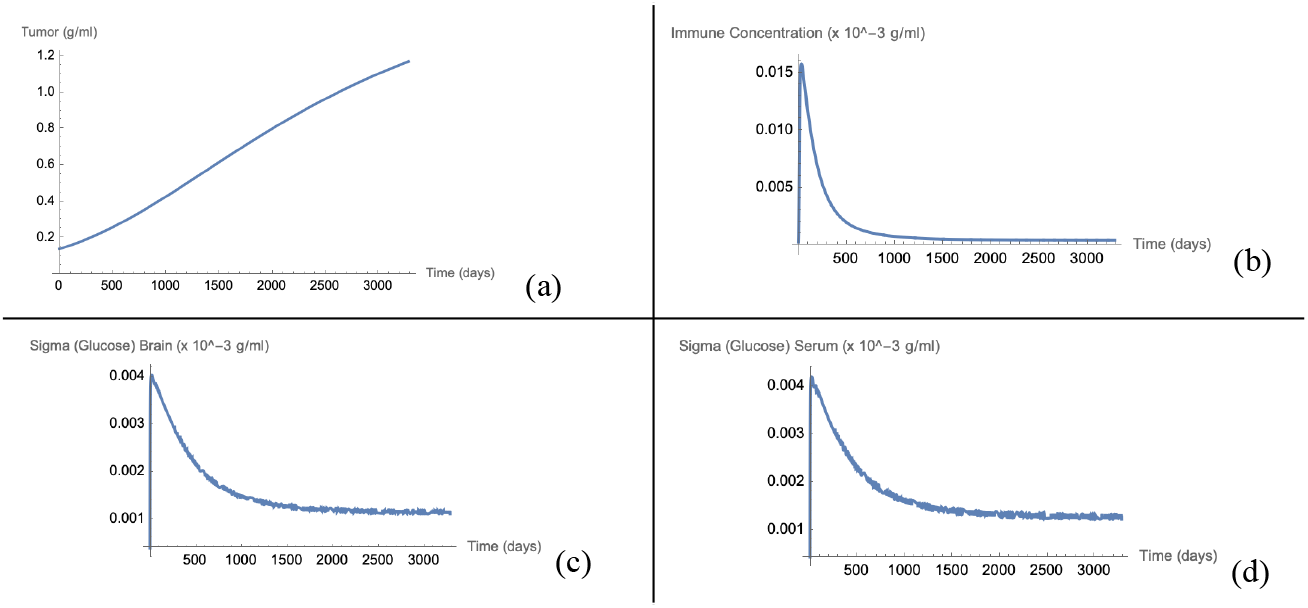
Steady state defined by Equation 6 is modeled to 9 years. Model is clearly not at equilibrium, with tumor neither at dormancy or at a large, aggressive state at which growth is zero. (a) Evolution of glioma, (b) Evolution of immune system activity, (c) Glucose levels in the brain, (d) Glucose levels in serum.

**Figure 2.**
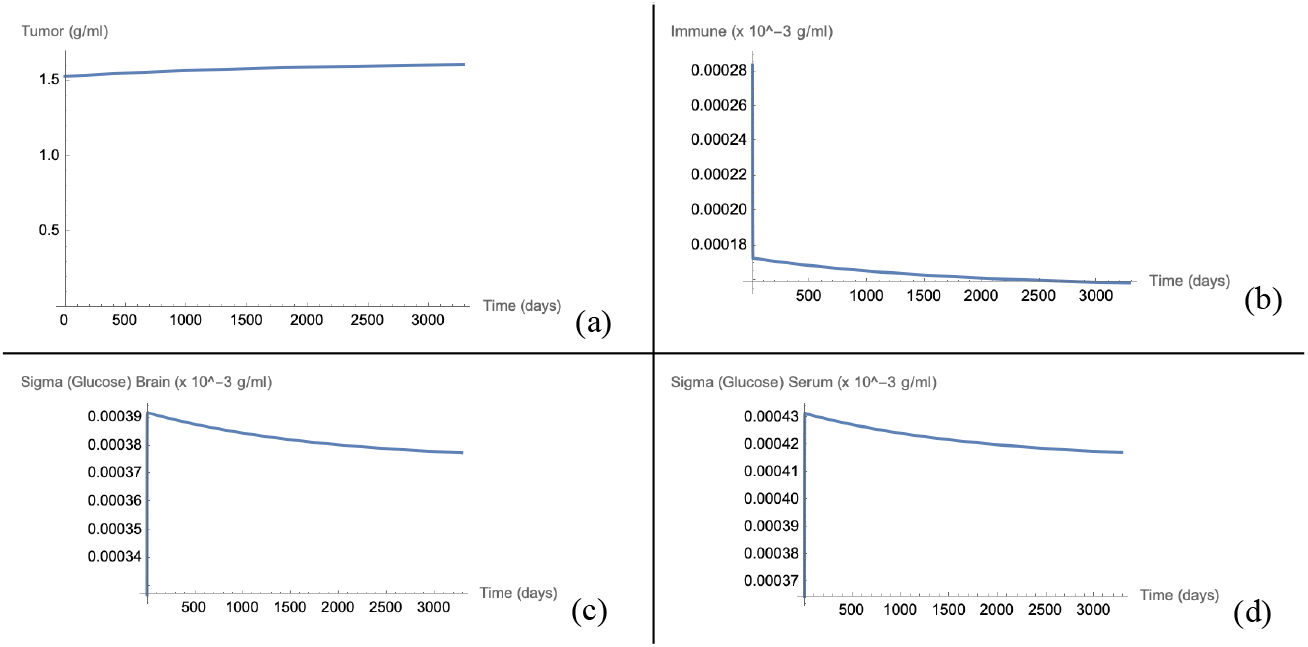
Steady state defined by Equation 9 is modeled to 9 years, displaying a system that is close to a steady value, although has significant spikes (∼20-50% of value) near time t = 0 as levels rapidly adjust to actual equilibrium. (a) Evolution of glioma, (b) Evolution of immune system activity, (c) Glucose levels in the brain, (d) Glucose levels in serum.

In this reproduction, Wolfram Mathematica 13.3.0’s NDSolve is used. Figures 1 and 2 illustrate that the steady states that Sturrock et al. solve for do not exactly quantitatively match those in the reproduction of their system shown above. This is in part due to the glucose intake function whose *Sin* function causes ODE solvers to propagate negative values at extremely small values, as well as the specific solver used. In order to prevent the latter from happening, for the sole purpose of identifying steady states, the glucose intake function (Equation 5) is fixed to σ_*min*_ . In order to find the steady states of this system, Mathematica’s NSolve is utilized, which has states that correspond to but do not exactly quantitatively match those of Sturrock et al.:

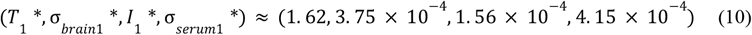

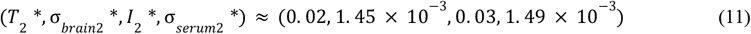

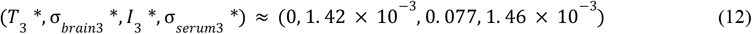

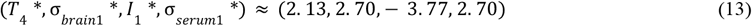

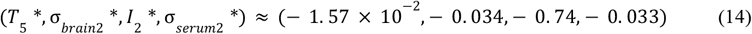

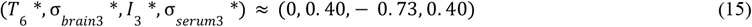

with Equation 10 representing the large, aggressive tumor and illustrated in Figure 3, Equation 11 representing the small, dormant tumor and illustrated in Figure 4, Equation 12 representing the healthy patient and illustrated in Figure 5, and Equations 13-15 representing the non-biologically relevant steady states.

**Figure 3.**
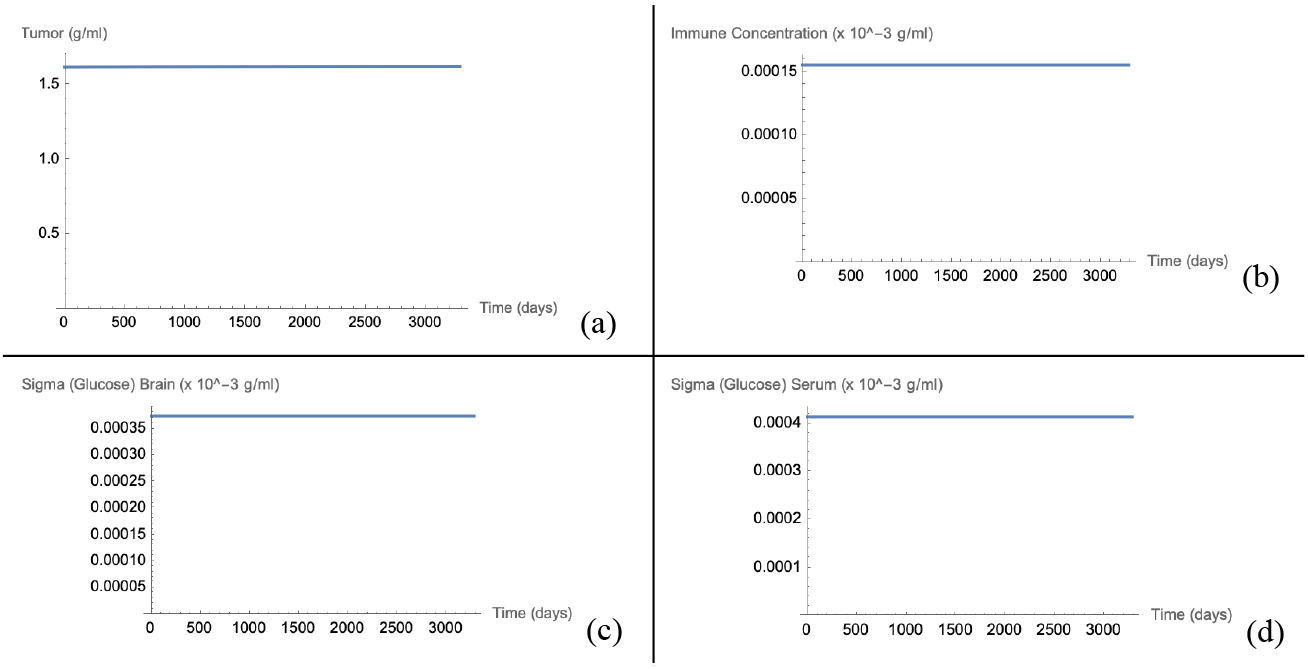
Steady state defined by Equation 10 is modeled to 9 years. (a) Evolution of glioma, (b) Evolution of immune system activity, (c) Glucose levels in the brain, (d) Glucose levels in serum.

**Figure 4.**
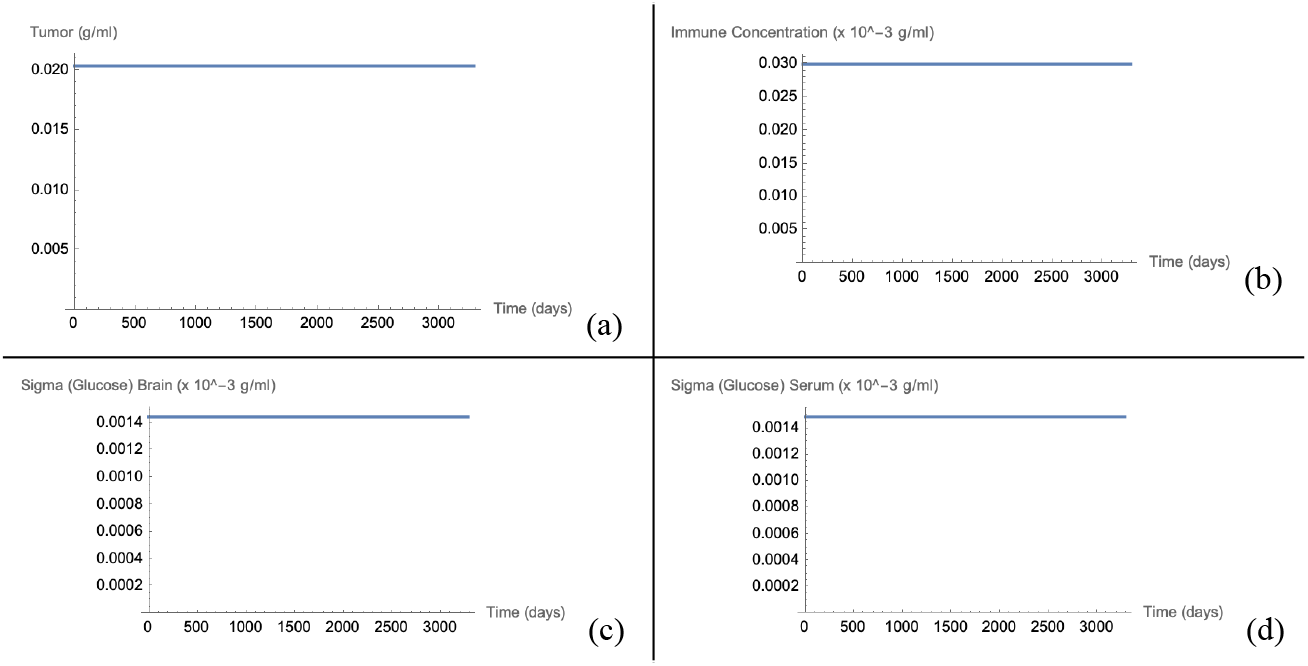
Steady state defined by Equation 11 is modeled to 9 years. (a) Evolution of glioma, (b) Evolution of immune system activity, (c) Glucose levels in the brain, (d) Glucose levels in serum.

**Figure 5.**
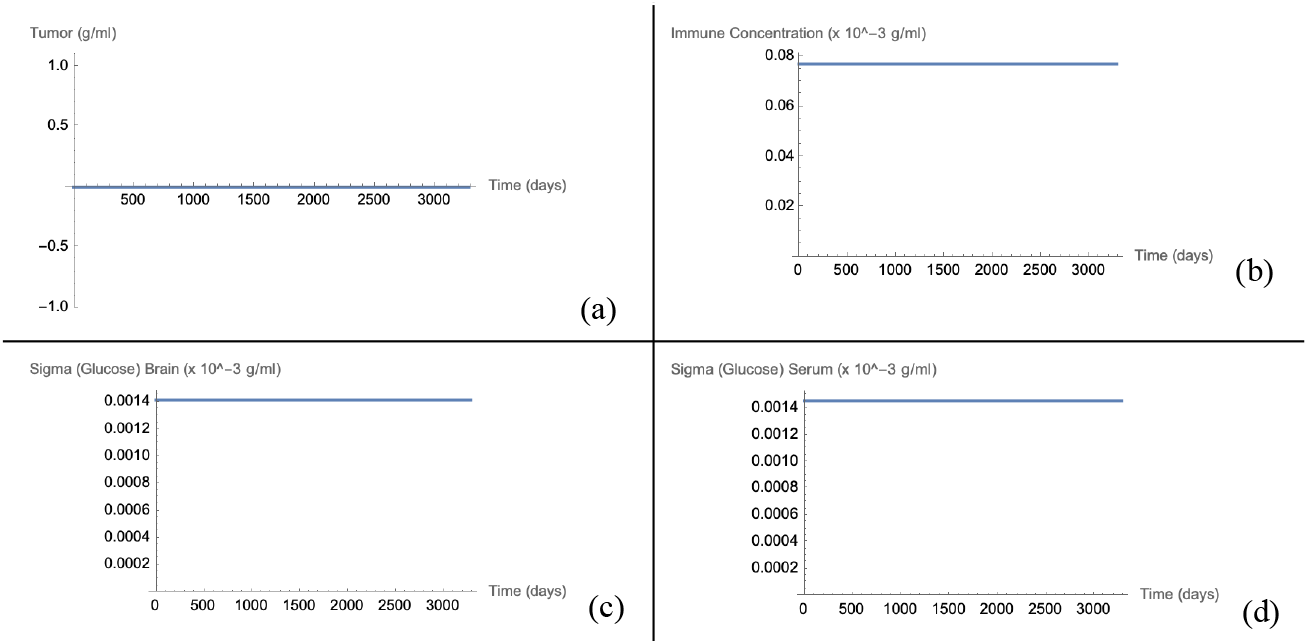
Steady state defined by Equation 12 is modeled to 9 years. (a) Evolution of glioma, (b) Evolution of immune system activity, (c) Glucose levels in the brain, (d) Glucose levels in serum.

### 2.3. Effect of Sudden Large Glioma Growth

There are physiologically relevant scenarios under which a patient’s serum glucose levels are substantially affected over a longer period of time. In particular, Sturrock et al. observe a scenario in which a previously dormant tumor (represented by Equations 7, 11) becomes stable as an aggressive glioma. This occurs when a patient adopts a glucose rich diet after 3 years, which affects the glucose intake function as shown in Equation 16:

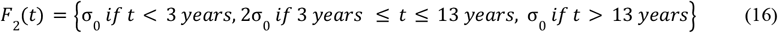

This glucose intake function is implemented, given the dormant tumor steady state as defined by Equation 11 and modeled as illustrated in Figure 6:

**Figure 6.**
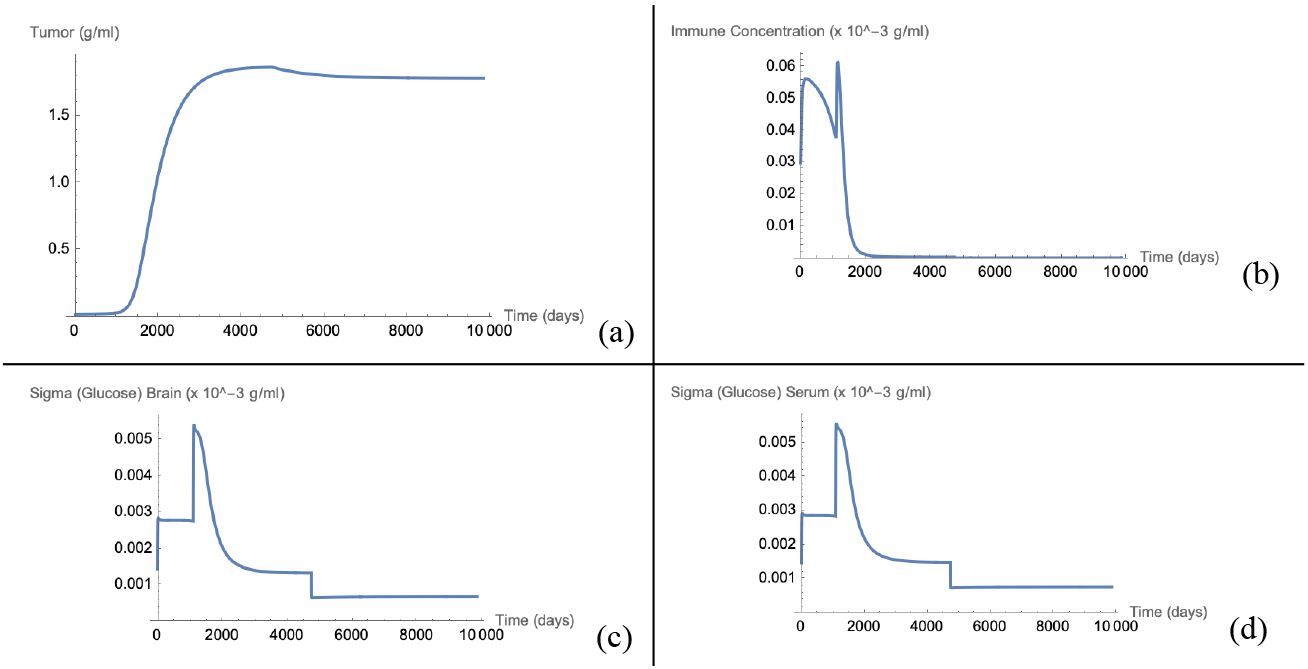
Steady state defined by Equation 11 is modeled to 27 years, given the new glucose intake function. (a) Evolution of glioma, (b) Evolution of immune system activity, (c) Glucose levels in the brain, (d) Glucose levels in serum.

When a glucose rich diet is adopted after 3 years, the tumor jumps from the unstable, dormant state to an aggressive, rapidly growing tumor. After 13 years, once the initial glucose intake is utilized again, the patient’s tumor slightly tapers off, as it is limited by glucose levels. However, the immune cell concentration never recovers as the tumor is too large. There are more interpretations of physiologically relevant glucose intake variations, which are explored in greater depth in this report’s revision of the Sturrock et al. model.

### 2.4. Glioma Cells Eradicating Immune Cells

The extent to which glioma cells can damage the immune system is important in pre-diagnostic gliomas, as the immune system is a natural inhibitor of these tumors. In the original Sturrock et al. model, this process is described as *d*_*TT*_*TI*, defining how the number of glioma cells eradicates the number of immune cells. Sturrock et al. found that the steady state (represented by Equation 11) corresponding to a dormant tumor illustrated a decaying effect as its ability to destroy immune cells increased—an unexpected effect. The Sturrock et al. reproduced system is simulated with varying values of *d*_*TT*_ in Figures 7-8 for the dormant tumor steady state:

**Figure 7.**
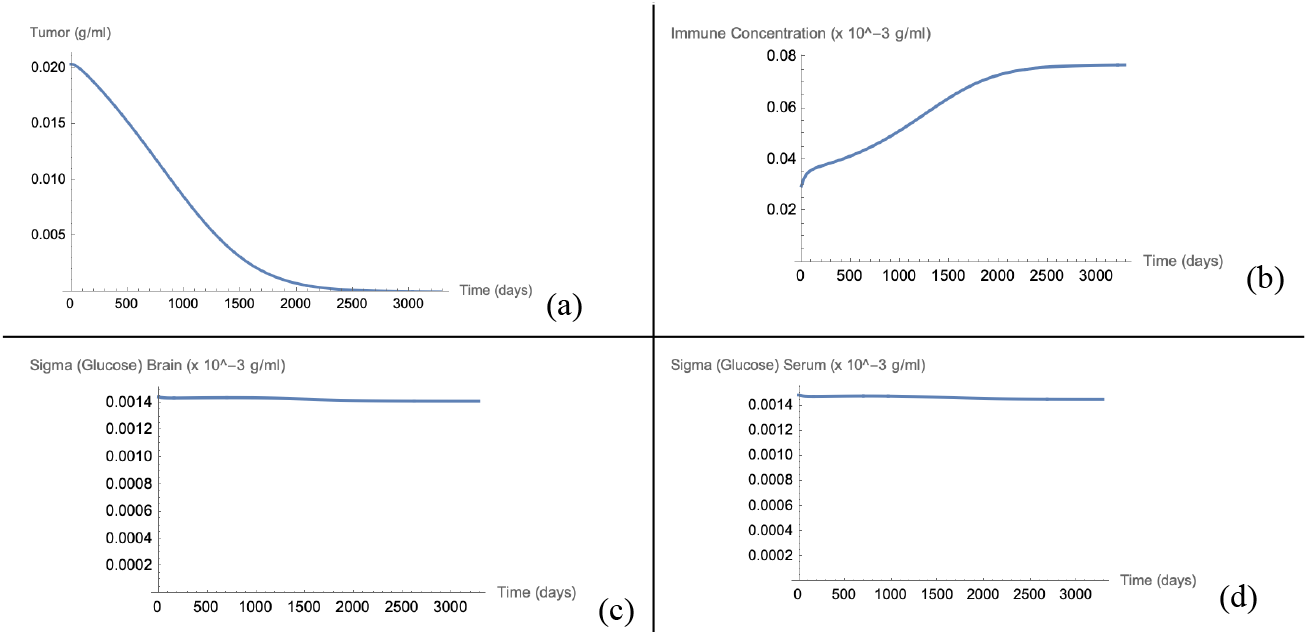
Steady state defined by Equation 11 is modeled to 9 years, given a d_TT_ value of 0.5 day ^−1^ . (a) Evolution of glioma, (b) Evolution of immune system activity, (c) Glucose levels in the brain, (d) Glucose levels in serum.

**Figure 8.**
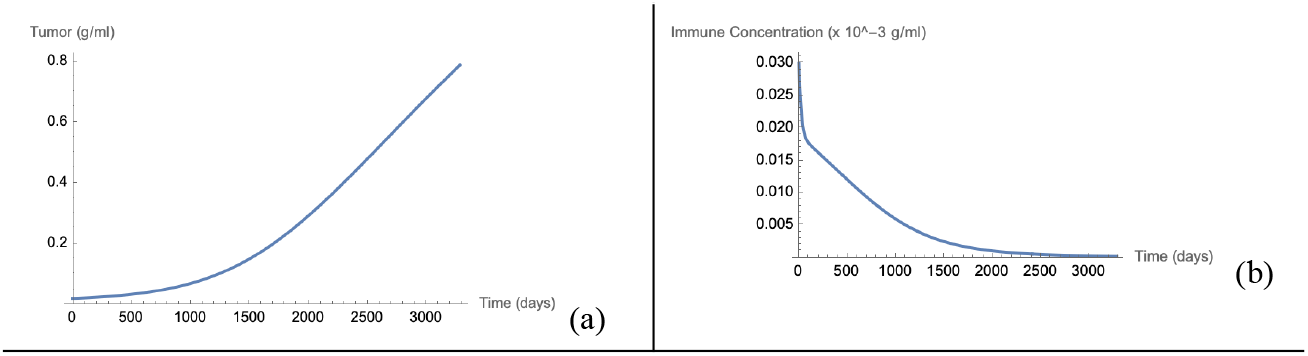
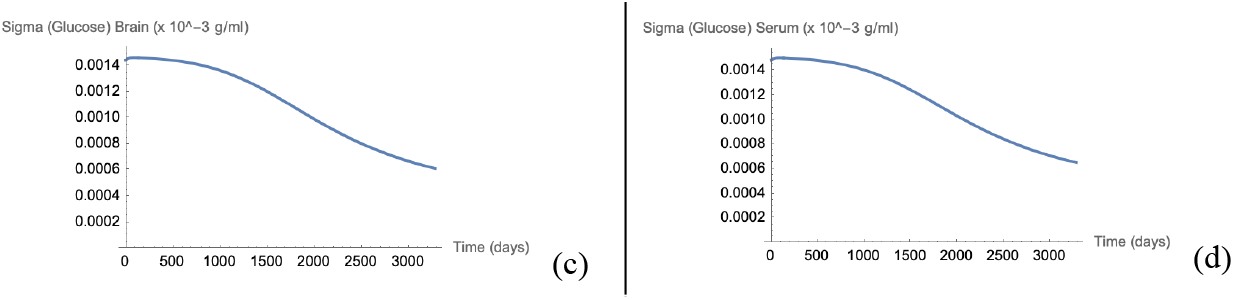
Steady state defined by Equation 11 is modeled to 9 years, given a d_TT_ value of 0.5 day^−1^ . (a) Evolution of glioma, (b) Evolution of immune system activity, (c) Glucose levels in the brain, (d) Glucose levels in serum.

Notably, when the parameter is increased to 1.5 *day* ^−1^, the tumor becomes more effective at killing immune cells, and as a result, shoots out of dormancy. When it is reduced to 0.5 *day*^−1^, the immune response is able to sufficiently fend it off. This differs from the Sturrock et al. results, even upon using the coarser glucose intake function, and the exact same steady state values. The behavior for the other steady states (i.e. aggressive and no tumor) are qualitatively as expected, corresponding to Sturrock et al.’s results. These can be found in Figure A1-A4.

## 3. Revising The Model

This report further develops the work of Sturrock et al. (2015) by revising their model in order to incorporate more biologically relevant parameters and processes. The revision includes introducing the concepts of vascularization and mutational burden, both of which are known to affect glioma growth and tumor-immune interactions. The revision is explored by varying model parameters pertaining to angiogenesis and immune system dynamics (3.1). Additionally, the original model is tested through further scenarios, simulating variable glucose inputs tuned to various orthogonal combinations of glucose intake timings and quantities (3.2).

### 3.1 Vascularization and Mutational Burden

Vascularization has a direct effect on tumor growth, with tumors needing to develop a network of vessels for efficient delivery of nutrients and oxygen. From an angiogenic point of view, vascularization of tumors is composed of two elements: the permeability of endothelial junctions and the rate of angiogenic sprouting (Yuan et al., 1996). The permeability of endothelial junctions depends on tumor contact area and the number of functional vessels, while the vascular factor is a representation of the fraction of vasculature that is exposed to tumor (George & Levine, 2020; Yuan et al., 1996; Herman et al., 2011). These processes can be represented by the amount of tumor cells that are sufficiently vascularized *v*, and parameters *ν*_*factor*_ and *ν*_*perm*_ representing the vascularization factor and vascular permeability, respectively. It is necessary for the tumor to have its vascular network established in order to invade and colonize regions that can further support its continued growth, as it otherwise is under a higher likelihood of perishing to immune response (Yuan et al., 1996).

The second major process introduced is mutational burden. Mutations are responsible for various alterations in the way cells respond to stimuli, potentially leading to tumor-immune subversion or escape (Wilkie et al., 2016). Mutations in tumor cells increase as they undergo clonal expansion, potentially allowing them to evade the immune system and therefore contribute to tumor growth. Wilkie et al. describe mutations as directly affecting the inverse immune response to the tumor growth. This revision assumes mutations accumulate both randomly over time and directly proportional to the number of tumor cells. The mutations can lead to more aggressive tumor behavior. In order to model the influence of mutations, a variable *M* is introduced to measure concentration of mutated tumor cells, while parameters *M*_*burden*_ and *M*_*rate*_ represent the proportion of tumor that has the propagated evasionary phenotype, and number of days to reach a mutation that achieves immune evasion, respectively.

The resultant system is given in equations 17-19 with equations 2-4 repeated for clarity:

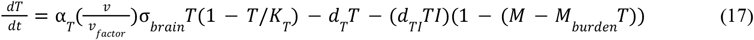

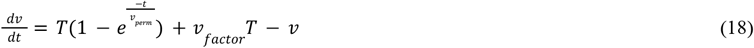

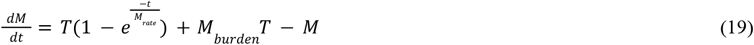

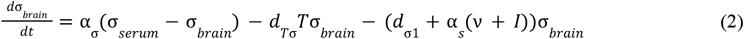

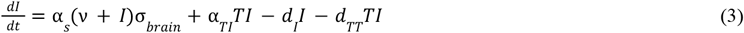

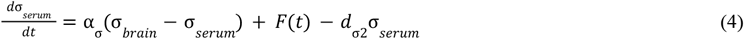

where the former parameters from Sturrock et al. can be found in Table 1 and newly introduced parameters in Table 2.

**Table 2.** Parameters for differential equation system, estimated and adapted from literature.

| Parameter | Description | Range (unit) | Source |
| --- | --- | --- | --- |
| $v_{factor}$ | Vascularization factor | 0.2 (none) | George & Levine (2020) |
| $v_{perm}$ | Vascular permeability due to formation of functional vascular network | 5000 (day) | Yuan et al. (1996) |
| $M_{burden}$ | Strength of immune response based on | 0.208 (none) | Wilkie et al. (2016) |
| $M_{rate}$ | Time to hit immune evasion mutation | 1270 (day) | Estimate |

The system has vascularization manifest at the start of equation 17, as a factor being multiplied by α _*T*_, thereby affecting its growth rate. Mutation burden affects the *d*_*TI*_ *TI* term such that the degradation of tumor cells caused directly by the immune system is dependent on the extent to which the tumor can evade immune response via mutations. The exponential term in Equations 18-19 is inspired by the cumulative distribution function of the Poisson distribution, where the probability of an event occurs in a fixed interval of time *t*.

The revised system is tested on the steady states defined in Equations 10-12, and the results are shown in Figures 9-11:

**Figure 9.**
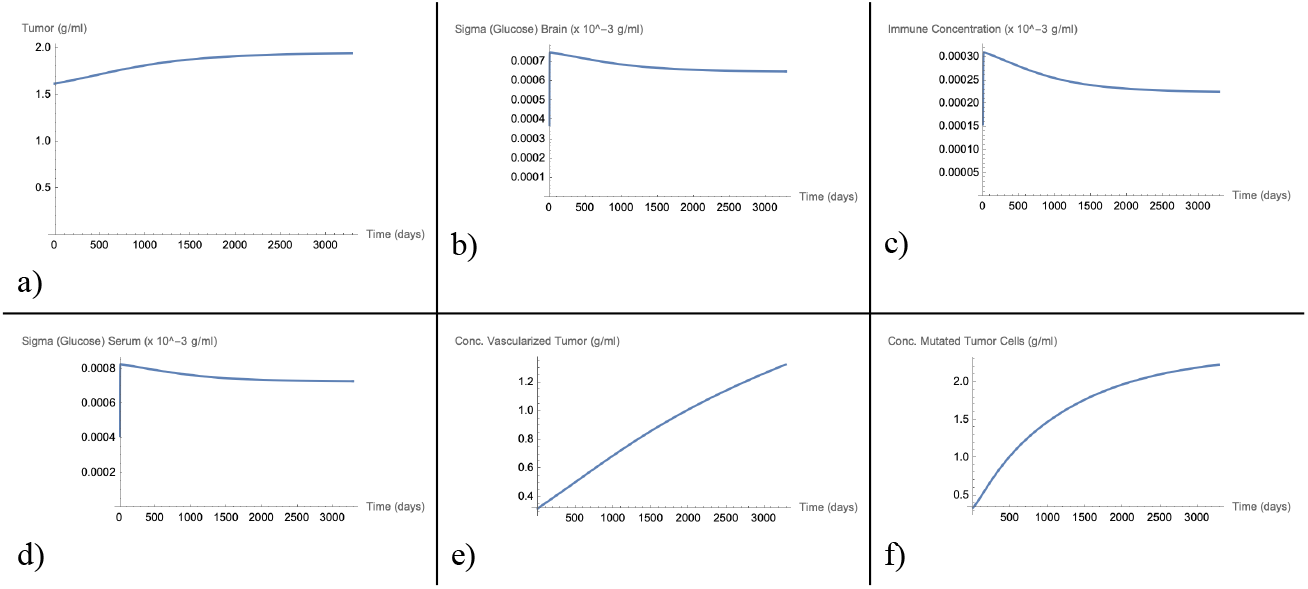
Aggressive tumor steady state (previously stable) is pushed towards carrying capacity as vascular network forms, and tumor accumulates sufficient mutations to evade immune concentration. (a) Evolution of glioma, (b) Glucose levels in the brain, (c) Evolution of immune system activity, (d) Glucose levels in serum, (e) Concentration of sufficiently vascularized tumor, (f) concentration of tumor that has immune-evasion.

**Figure 10.**
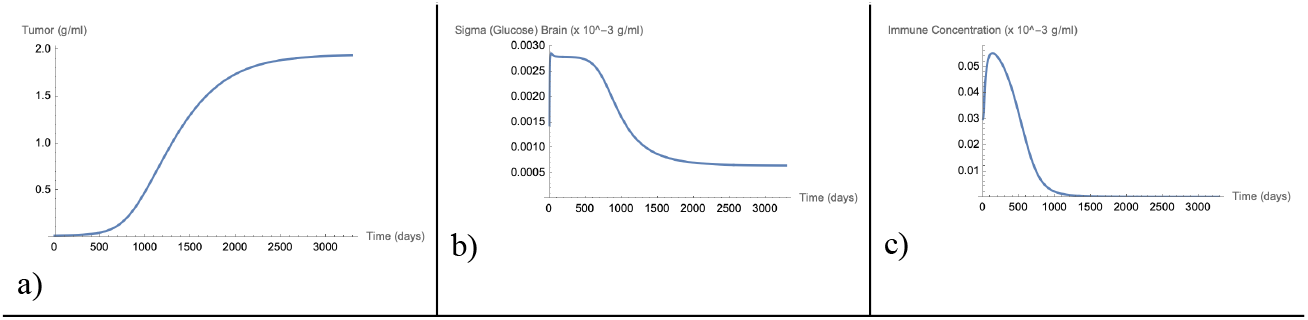
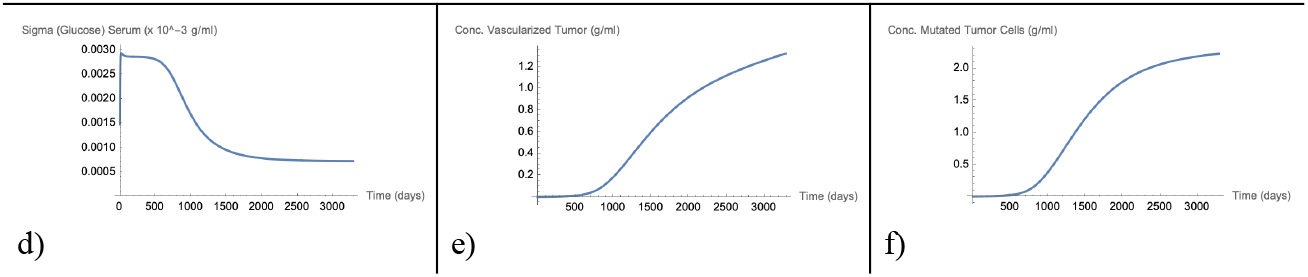
Dormant tumor steady state (unstable) is pushed towards carrying capacity as vascular network forms, and tumor accumulates sufficient mutations to evade immune concentration. (a) Evolution of glioma, (b) Glucose levels in the brain, (c) Evolution of immune system activity, (d) Glucose levels in serum, (e) Concentration of sufficiently vascularized tumor, (f) concentration of tumor that has immune-evasion.

**Figure 11.**
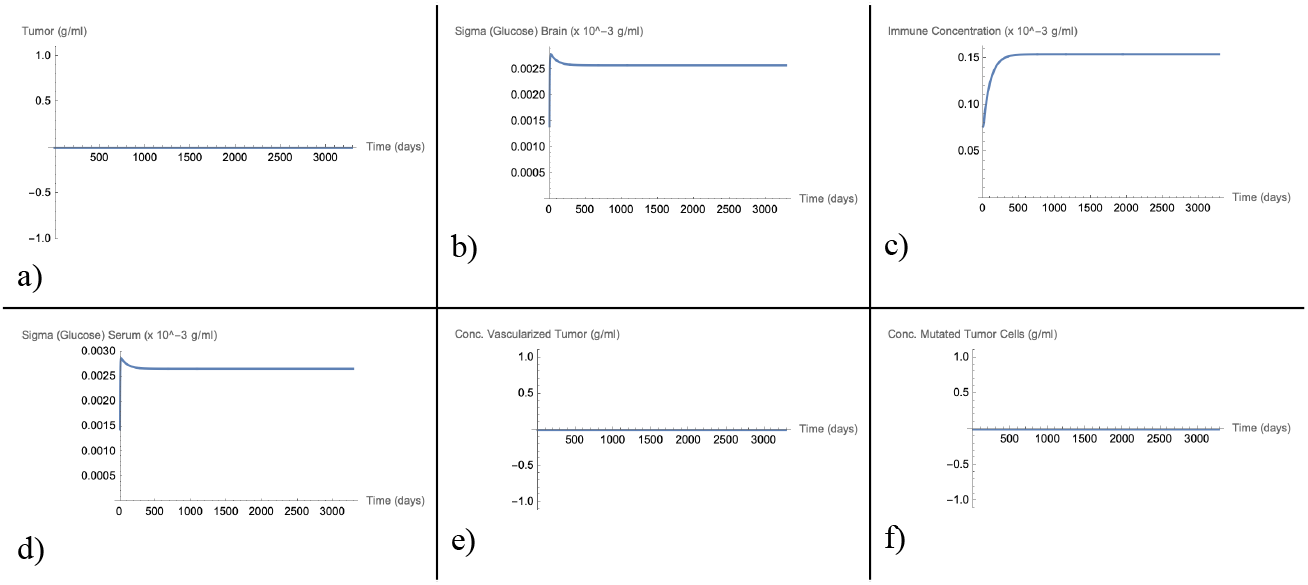
No tumor steady state (healthy) pursues no vascular network, and no accumulation of mutations. (a) Evolution of glioma, (b) Glucose levels in the brain, (c) Evolution of immune system activity, (d) Glucose levels in serum, (e) Concentration of sufficiently vascularized tumor, (f) concentration of tumor that has immune-evasion.

### 3.2. Glucose Intake Variability

In addition to a patient adapting a glucose rich diet, it is very possible for the opposite to occur. In fact, conditions such as sarcopenia may influence a patient’s diet substantially, causing a drop in glucose intake (Zhao et al., 2015). Although these tumors are prediagnostic, there is an interest in understanding how a tumor’s behavior responds to a drop in glucose intake as this could also pertain to a patient getting weaker or aging. The resultant glucose intake function *F(t)* is defined in Equation 20:

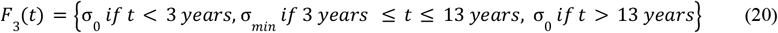

where *t* is a positive number in days. Glucose intake starts at σ_0_, becoming σ_*min*_ after 3 years, and back to σ_0_ after 13 years. This may represent a scenario where a patient has a glucose-limiting diet for an extended period of time. The original system with this glucose intake function is tested on the steady states defined in Equations 10-12, and the results are shown in Figures 12-14:

**Figure 12.**
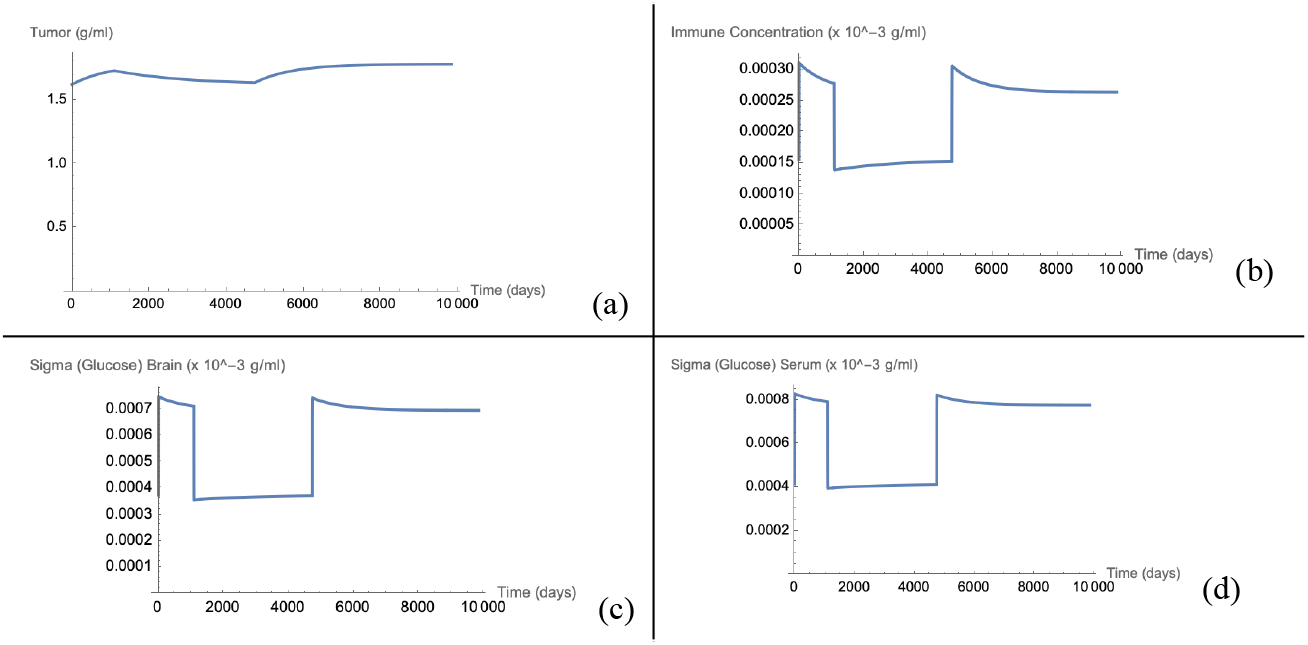
Aggressive tumor (steady state defined by Equation 10) is modeled to 27 years, given the new glucose intake function. (a) Evolution of glioma, (b) Evolution of immune system activity, (c) Glucose levels in the brain, (d) Glucose levels in serum.

**Figure 13.**
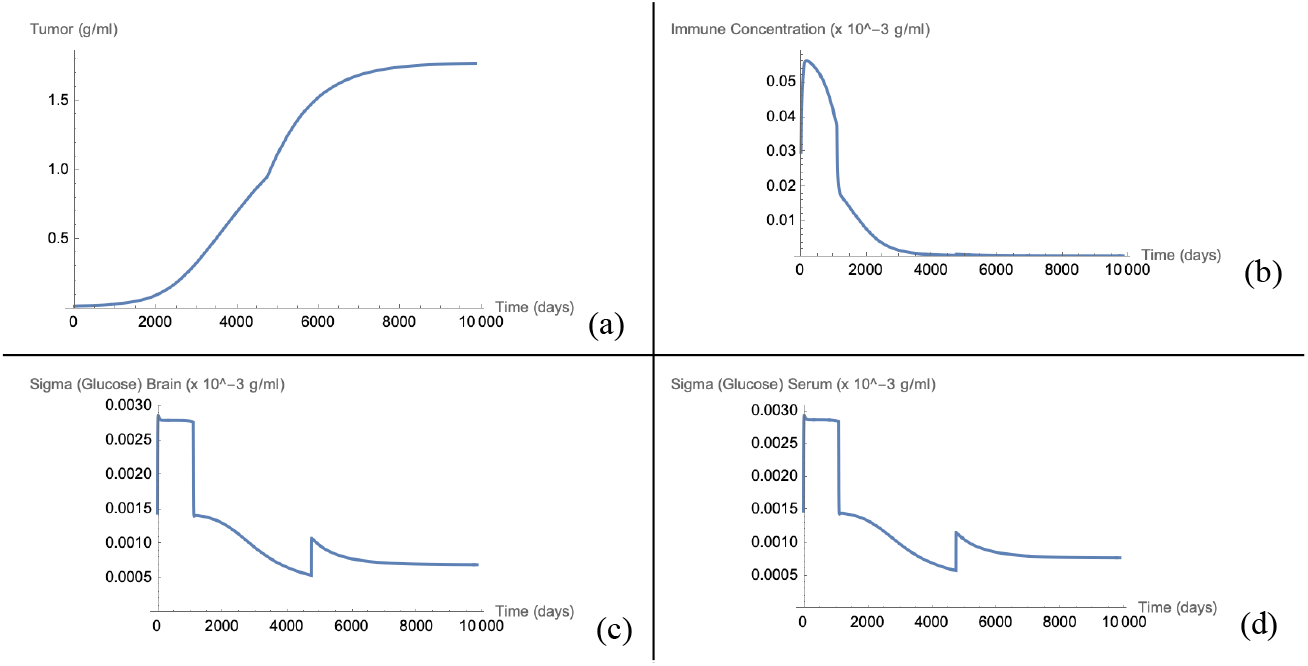
Dormant tumor (steady state defined by Equation 11) is modeled to 27 years, given the new glucose intake function. (a) Evolution of glioma, (b) Evolution of immune system activity, (c) Glucose levels in the brain, (d) Glucose levels in serum.

**Figure 14.**
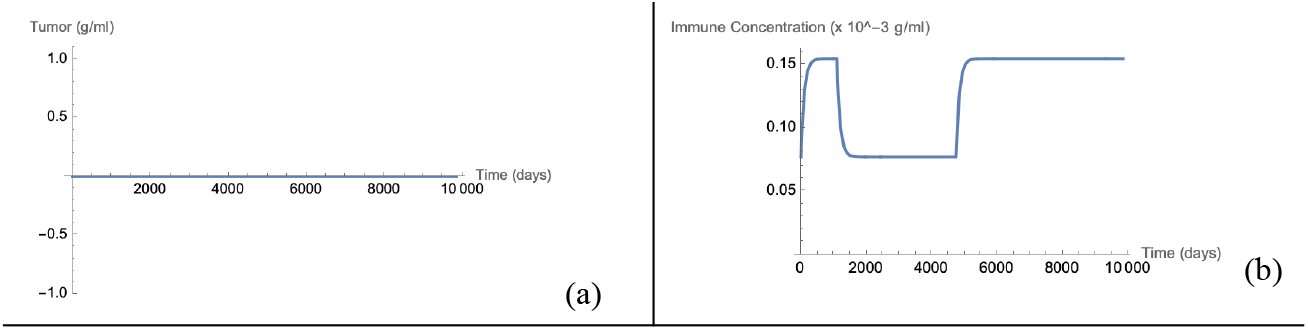
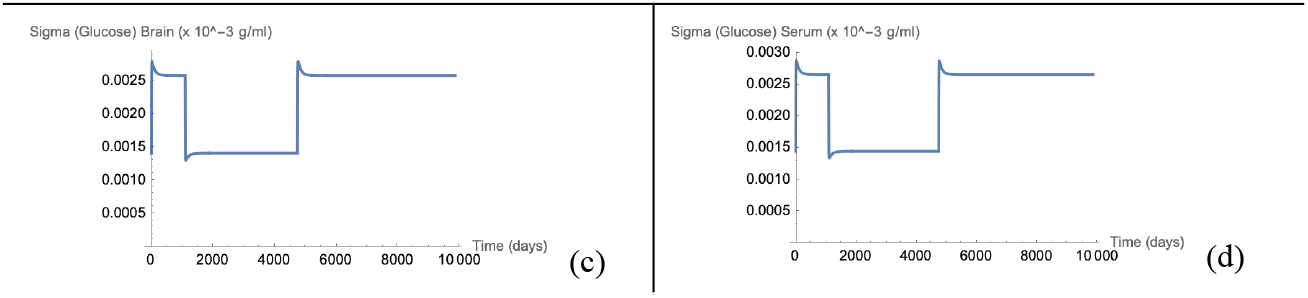
No tumor (steady state defined by Equation 12) is modeled to 27 years, given the new glucose intake function. (a) Evolution of glioma, (b) Evolution of immune system activity, (c) Glucose levels in the brain, (d) Glucose levels in serum.

## 4. Discussion

This report reproduced the major results of the Sturrock et al. (2015) mathematical model for glioma-glucose-immune interactions, including the three relevant steady states. The focus of the reproduction was on how the variables interact and contribute to steadiness of the system, and further insights into the behavior of pre-diagnostic gliomas. During the reproduction of their results, several limitations with software and modeling were identified. Due to the nature of the glucose intake function *F(t)* utilizing a *Sin* and *Max* function, identifying the steady states of the model was difficult. Simplifications had to be made — including modifying F(t) itself for the purpose of modeling. As *t* increased, *F(t)* often caused the differential equation solver to return negative values for glucose intake, thus reducing the ability of the solver to identify the steady states correctly. Adding a constant to force the differential equation solver to propagate non-negative values for glucose intake during reproduction did not perfectly return Sturrock et al. values, thus the model’s steady states were only qualitatively validated. It was also impossible to specify initial conditions to the model that exactly align with the solved steady states—thus leading to extremely minor variations in variables over huge domains of time. Thus, most of the figures presented in this report have had their axes tweaked to minimize this effect.

In section 2.3, when the effect of sudden large glioma growth is modeled using *F2(t)*, a substantial result emerges, showcasing a limitation of the Sturrock et al. model. The glucose-rich diet that manifests after 3 years causes glucose levels to spike, leading to an increase in tumor growth that allows the tumor to escape its dormant state and become aggressive. This substantial growth causes a death in immune cells that appears to be unrecoverable. Later, once the initial diet/intake is back to normal, the tumor barely decays. This is because the immune activity is still near zero. While an interesting circumstance theoretically, this may not be physiologically accurate. It is possible that the tumor still experiences immune pressure, as the immune system regains potency. This limitation can potentially be overcome by introducing a noise-generating functionality in the immune activity in order to allow more dynamic behaviors. However, as seen in this report’s revised model, with the addition of an immune evasion variable, it is expected that the glioma will be able to hide from circulating immune cells and, as shown, remain large in size.

This does create interest in an opposite scenario that discusses the opposite of a glucose-rich diet. In section 3.2, *F3(t)* simulates this very scenario, demonstrating that the tumor decays, albeit only slightly in the aggressive steady state, and there is a minimal effect on the tumor growing to full capacity from the dormant steady state. If anything, the limited glucose diet has more of a role in immune activity, which is why the tumor proceeds to grow without much recoil. For the Sturrock et al. system overall, the high tumor growth rate almost always results in the tumor exceeding some threshold that would keep it quiescent. In other words, most tumors—once established—continue to grow until they reach their carrying capacity. This illustrates the deadliness of gliomas, however, it appears that the tumor continues to grow or be sustained, even if glucose levels are near zero—something that may not be biologically possible. The model also fails to capture the competition between other brain cells and the glioma, something that requires further inquiry in future mathematical models.

In the other revisions of the model, where vascularization and immune evasion are introduced, several conclusions can be drawn from the results. The three revised model scenarios described in section 3.1 show how the model structure can be manipulated to study different scenarios regarding tumor-immune interactions. The results suggest that the introduction of the parameters simulating vascularization and mutational burden gives a better indication of how glioma tumors may interact with the immune system. In particular, the new system illustrates the dependence of the tumor—especially for its dormant steady state—on vascularization quantities and mutation rate. The mutation rate had quite a substantial effect, allowing the dormant stage to proceed to aggressive growth upon accumulating its immune-invasive mutation, and this mutation became highly prevalent in the tumor mass. The Poisson distribution with time dependence is introduced in both differential equations because it helps elucidate the probability that an immune-evasive mutation arises (event) occurring a certain number of times in *t*. Future research into parameter influence on the system for this model would be extremely beneficial. For instance, decreasing tumor size with low *ν*_*factor*_ and or introducing a tumor with a lower *M*_*rate*_ speaks to the heterogeneous nature of tumors, and how patients may have different progression speeds of tumors depending on the tumor environment as well as the mutations in the tumor cells—and could have implications for suggesting personalized medical treatments as opposed to cocktail therapies for patients with gliomas.

The applications of these findings are several. Clinical therapies for gliomas are often harsh and lack a proper understanding of tumor-immune interactions. Mathematical modeling clearly outlines how multiple mechanisms interact to create or suppress tumor growth, helping clinicians better understand pre-diagnostic gliomas. The understanding of tumor-immune dynamics could also help with patient survivability—possibly allowing clinicians to detect tumors at early stages. Sturrock et al. focus on screening and prevention for early detection of gliomas—and while mathematically simpler, the Sturrock et al. model provides evidence into how venous permeability of endothelial junctions as well as the rate of angiogenic sprouting could aid screening efforts. Physiologically, the revision of the Sturrock et al. model paves a more detailed path for understanding the growth (or lack thereof) of pre-diagnostic gliomas—which can better explain patient treatments. Additionally, the incorporation of mutational burden in the system showcases the malignant nature of gliomas. In terms of glucose intake, this could have implications for proper nutrition for glioma patients—agents that adopt a glucose-rich diet should be alarming, as this could push the tumor into becoming aggressive. All these applications necessitate further development not only of the methodology presented here but of further mathematical models that can simulate more clinically and biologically relevant phenomena.

## Data Availability

All data produced in the present study are available upon reasonable request to the authors

## 6. Appendices

**Figure A1.**
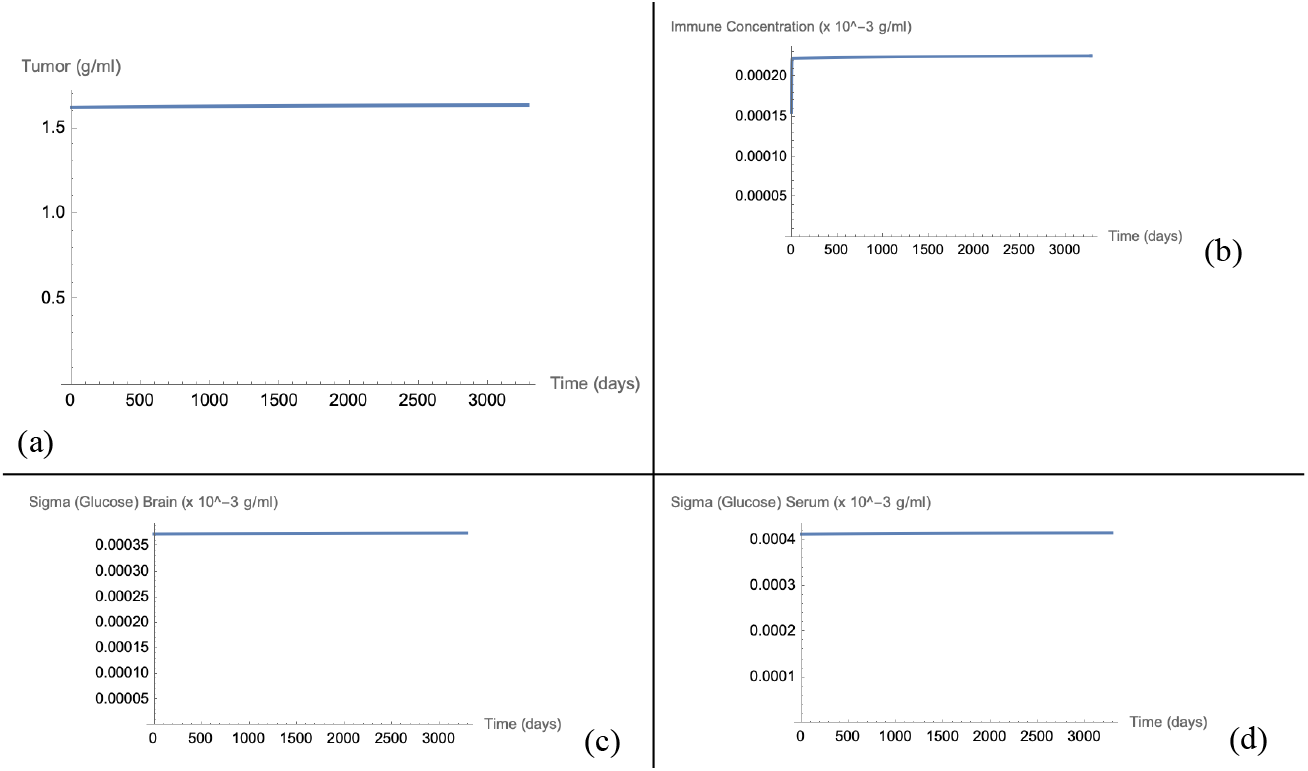
Steady state defined by Equation 10 is modeled to 9 years, given a d_TT_ value of 0.5 day^−1^. (a) Evolution of glioma, (b) Evolution of immune system activity, (c) Glucose levels in the brain, (d) Glucose levels in serum.

**Figure A2.**
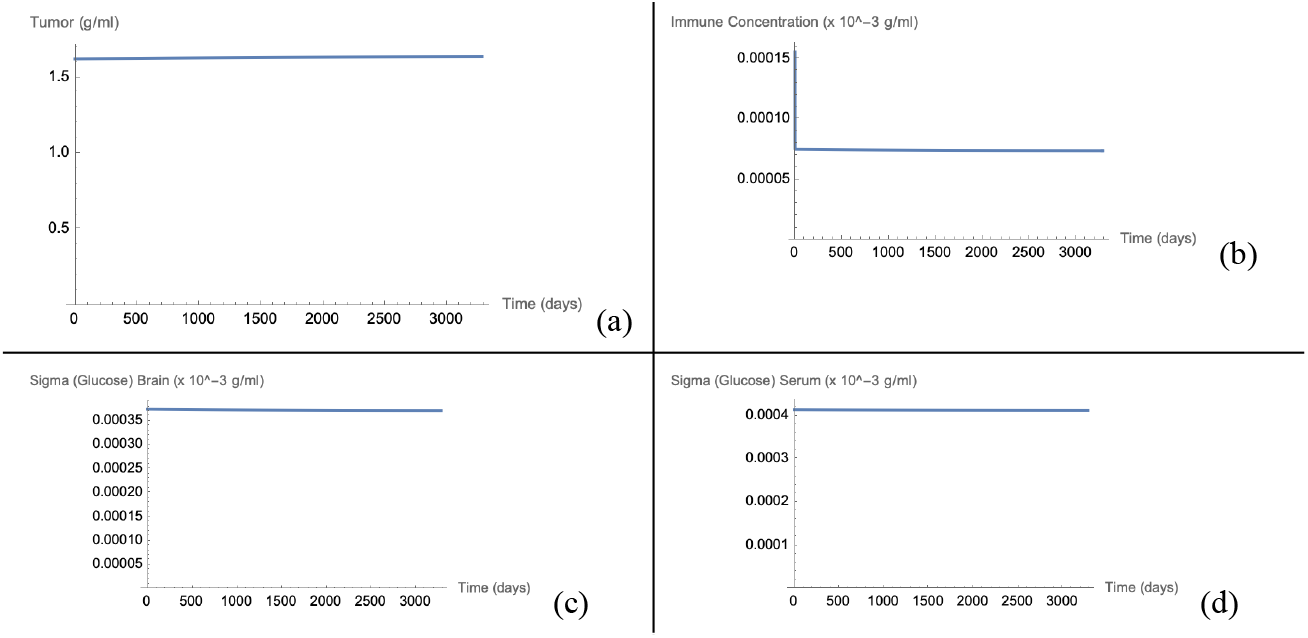
Steady state defined by Equation 10 is modeled to 9 years, given a d_TT_ value of 0. 5 day ^−1^. (a) Evolution of glioma, (b) Evolution of immune system activity, (c) Glucose levels in the brain, (d) Glucose levels in serum.

**Figure A3.**
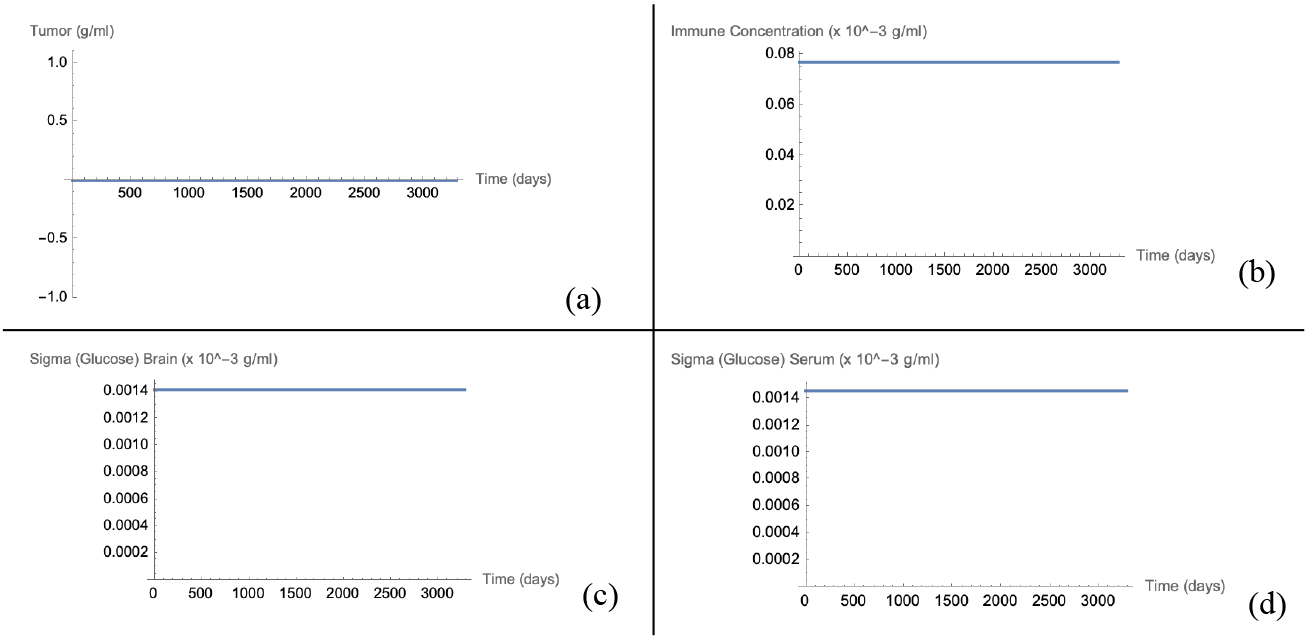
Steady state defined by Equation 12 is modeled to 9 years, given a d TT value of 1. 5 day ^−1^. (a) Evolution of glioma, (b) Evolution of immune system activity, (c) Glucose levels in the brain, (d) Glucose levels in serum.

**Figure A4.**
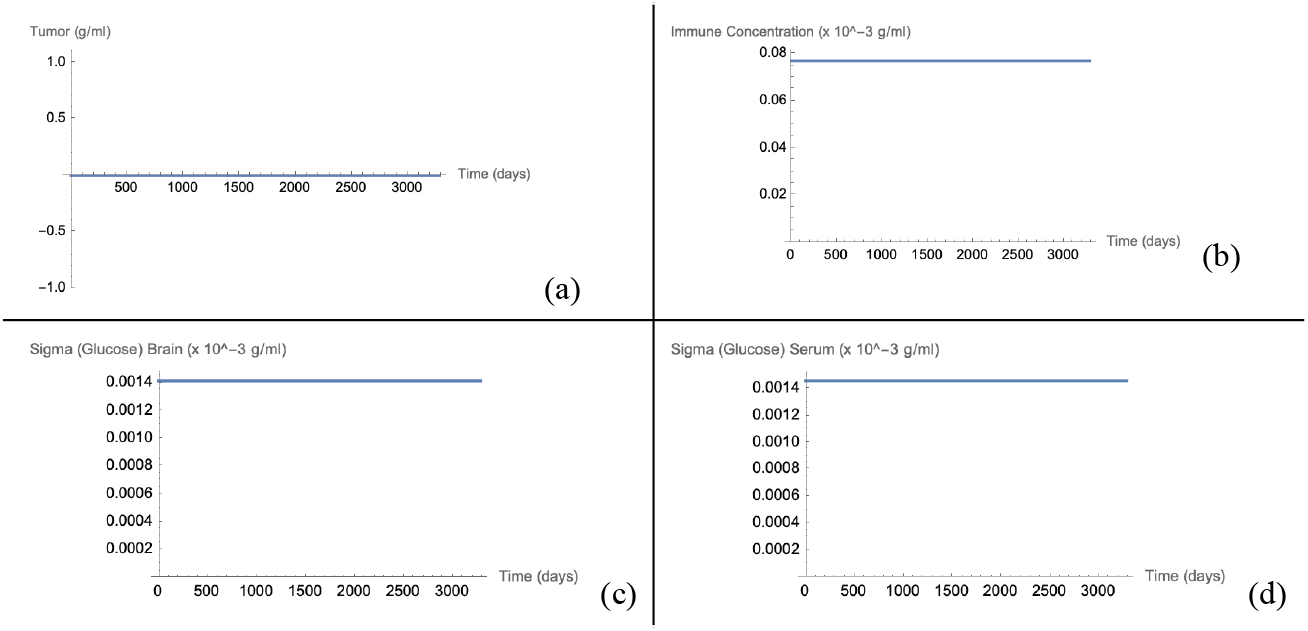
Steady state defined by Equation 12 is modeled to 9 years, given a d_TT_ value of 1. 5 day^−1^ . (a) Evolution of glioma, (b) Evolution of immune system activity, (c) Glucose levels in the brain, (d) Glucose levels in serum.

## Footnotes

1 The term steady states is used in place of equilibria as Sturrock et al. have elected its usage.

